# Bacteriophage and antibiotic combination therapy for recurrent *Enterococcus faecium* bacteremia

**DOI:** 10.1101/2022.11.23.22282657

**Authors:** Madison E. Stellfox, Carolyn Fernandes, Ryan K. Shields, Ghady Haidar, Kailey Hughes Kramer, Emily Dembinski, Mihnea R. Mangalea, Gregory S. Canfield, Breck A. Duerkop, Daria Van Tyne

**Affiliations:** Division of Infectious Diseases, Department of Medicine, University of Pittsburgh, Pittsburgh, Pennsylvania, USA; Department of Immunology and Microbiology, University of Colorado School of Medicine, Aurora, Colorado, USA

**Author notes:** Address correspondence to: Daria Van Tyne. Department of Microbiology, New York University Medical Center, New York, NY, USA. Goldbelt C6, Division of Healthcare Quality Promotion, Centers for Disease Control and Prevention, Atlanta, Georgia, USA.

**Keywords:** Bacteriophage therapy, vancomycin-resistant enterococci, microbiome

## Abstract

*Enterococcus faecium* is a member of the human gastrointestinal (GI) tract microbiota but can also cause invasive infections, especially in immunocompromised hosts. Enterococci display intrinsic resistance to many antibiotics, and most clinical *E. faecium* isolates have acquired vancomycin resistance, leaving clinicians with a limited repertoire of effective antibiotics. As such, vancomycin-resistant *E. faecium* (VREfm) has become an increasingly difficult to treat nosocomial pathogen that is often associated with treatment failure and recurrent infections. We followed a patient with recurrent *E. faecium* bloodstream infections (BSIs) of increasing severity that ultimately became unresponsive to antibiotic combination therapy over the course of 7 years. Whole genome sequencing (WGS) showed that the patient was colonized with closely related *E. faecium* strains for at least two years, and that invasive isolates likely emerged from a large *E. faecium* population in the patient’s GI tract. The addition of bacteriophage (phage) therapy to the patient’s antimicrobial regimen was associated with several months of clinical improvement and reduced intestinal burden of VRE and *E. faecium*. Eventual recurrence of *E. faecium* BSI was not associated with the development of antibiotic or phage resistance in post-treatment isolates. However, an anti-phage neutralizing antibody response occurred simultaneously with an increased relative abundance of VRE in the GI tract, both of which may have contributed to clinical failure. Taken together, these findings highlight the potential utility and limitations of phage therapy to treat antibiotic-resistant enterococcal infections.

**Importance:** Phage therapy is an emerging therapeutic approach for treating bacterial infections that do not respond to traditional antibiotics. The addition of phage therapy to systemic antibiotics to treat a patient with recurrent *E. faecium* infections that were non-responsive to antibiotics alone resulted in fewer hospitalizations and improved the patient’s quality of life. Combination phage and antibiotic therapy reduced *E. faecium* and VRE abundance in the patient’s stool. Eventually an anti-phage antibody response emerged that was able to neutralize phage activity, which may have limited clinical efficacy. This study demonstrates the potential of phages as an additional option in the antimicrobial toolbox for treating invasive enterococcal infections and highlights the need for further investigation to ensure phage therapy can be deployed for maximum clinical benefit.

## Observation

Enterococci are hardy and adaptable gastrointestinal (GI) tract commensal organisms found in almost all terrestrial animals (1). Enterococci display intrinsic resistance to many antibiotics, and most clinical *Enterococcus faecium* isolates in the United States have acquired vancomycin resistance, leaving clinicians with a limited repertoire of effective antibiotics. These characteristics have allowed enterococci to become successful nosocomial pathogens, especially in immunocompromised populations (2). Bacteriophages (phages) offer a potential adjunctive tool to manage infections with multi-drug resistant bacteria (3). However, few reports have demonstrated *in vivo* efficacy of phage therapy for *E. faecium* infections in humans (4). Knowledge gaps remain regarding the effects of phage therapy on GI tract microbiota and whether host immune responses complicate phage therapy. Here, we present an in-depth characterization of the clinical, microbiological, and host immune response aspects of phage therapy in a patient with several years of recurrent VRE bacteremia.

The patient was a female in her 50s with a past medical history significant for prior Roux-En-Y bariatric surgery, recurrent extended-spectrum beta-lactamase (ESBL)-producing gram-negative urinary tract and pulmonary infections, and mild immunosuppression secondary to treatment for Sjogren’s syndrome and adrenal insufficiency. She was known to the University of Pittsburgh Medical Center (UPMC) for recurrent *E. faecium* bloodstream infections (BSI) beginning in 2012. She experienced several hospitalizations for *E. faecium* BSI between 2012 and 2020 and received multiple courses of therapeutic and suppressive antibiotics. No focal source of infection was discovered despite extensive diagnostic imaging, including multiple transthoracic and transesophageal echocardiography examinations, tagged white blood cell and PET/CT scans, as well as endoscopy and colonoscopy procedures. Therefore, the patient’s recurrent bacteremia was attributed to reseeding from her GI tract microbiota. Starting in June 2020 (observation day 1), the patient experienced increased frequency and duration of *E. faecium* BSI events (Fig. 1A). This culminated in 26 days of persistent *E. faecium* bacteremia despite treatment with multiple antibiotics displaying *in vitro* activity (Event D, Fig. 1A). The patient was subsequently referred for salvage phage therapy.

**Figure 1.**
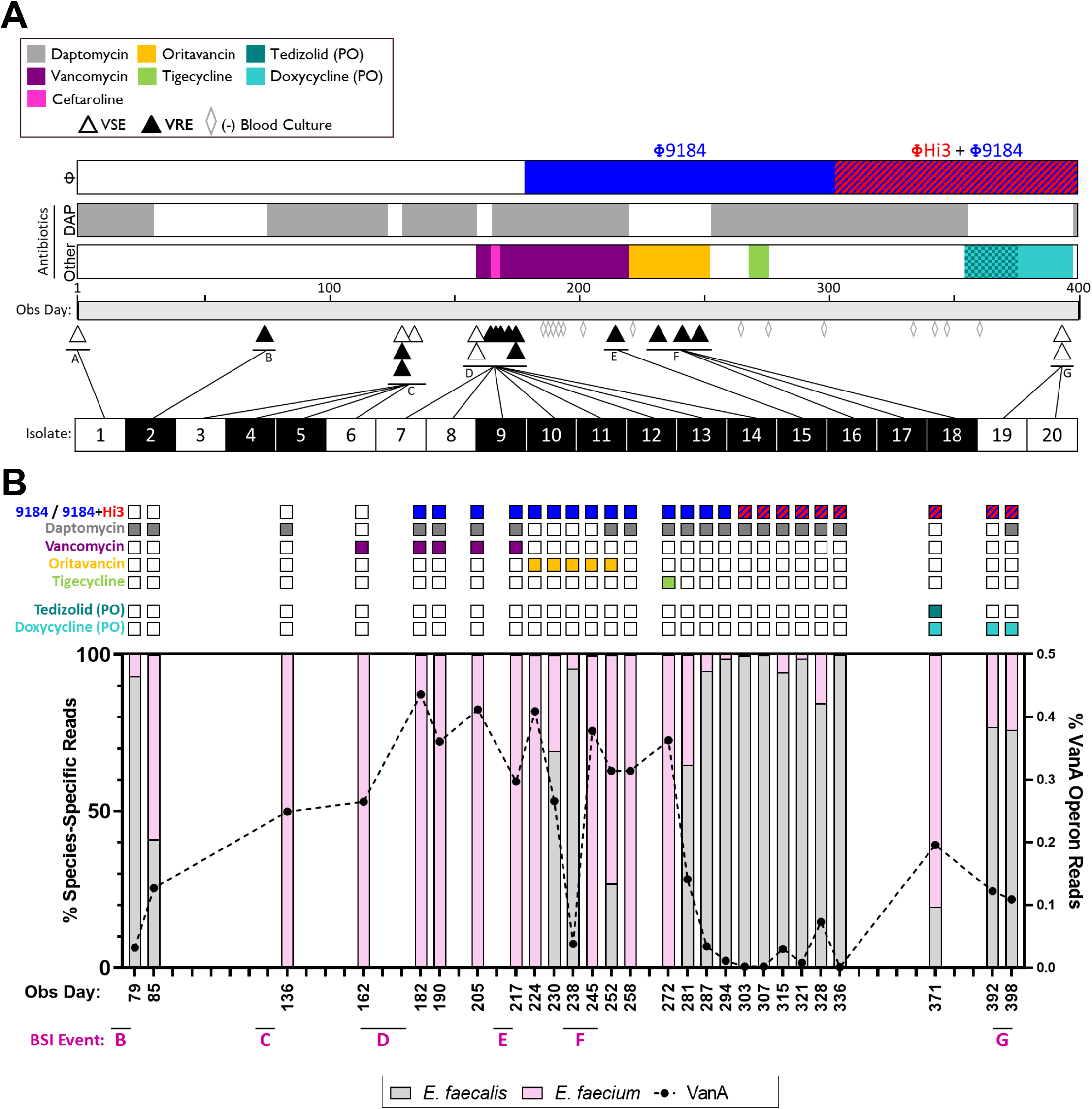
Clinical timeline and GI tract enterococcal populations. (A) Timeline represents the observation period (June 2020 through July 2021). The 3 bars above the timeline depict the type and duration of the treatments used throughout the observation period: blue (Φ9184) and red (ΦHi3) for phages, gray for daptomycin exposure, and the multicolored bar highlights the other systemic antibiotics used (vancomycin – purple, ceftaroline – pink, oritavancin – yellow, tigecycline – green, tedizolid – dark teal, doxycycline – light teal). PO indicates that the antibiotic was given orally. Triangles below the timeline indicate all positive blood cultures that grew vancomycin-sensitive *E. faecium* (VSE, open triangles) and vancomycin-resistant *E. faecium* (VRE, closed triangles). Gray, open diamonds indicate blood cultures that were obtained after starting phage therapy but remained negative. Below the timeline each BSI isolate that underwent WGS is numbered 1 through 20, shaded white for VSE and black for VRE. (B) GI enterococcal population metagenomics of pooled colonies from stool samples plated on bile esculin azide agar. Bar graph shows the relative abundance of reads mapping to *E. faecalis* (white bars), *E. faecium* (pink bars) and the VanA operon (black dotted line). Colored squares above each bar indicate which phage (top row), systemic intravenous antibiotics (rows 2 – 5) and oral antibiotics (rows 6 – 7) the patient was being treated with at the time the stool sample was collected.

A previously characterized siphovirus phage, Φ9184, had lytic activity against clinical isolates collected from the patient (5). After obtaining emergency investigational new drug (eIND) approval by the FDA, local IRB approval and informed patient consent, Φ9184 was added to systemic antibiotics on observation day 182 at 1×10^9^ plaque forming units (PFU) per dose. The phage formulation was given three times daily both intravenously (IV) and orally (PO) in an effort to target the suspected source of the recurrent infections, the GI tract. To prevent degradation of the phage in the stomach, the patient was maintained on a proton pump inhibitor regimen. After 26 days of persistent infection, bacteremia resolved within 24 hours of phage administration, and the patient was discharged home soon afterwards on antibiotic therapy (which alone had failed to eradicate the infection) and phage therapy (Fig. 1A).

While receiving concurrent treatment with daptomycin, vancomycin and Φ9184, a single outpatient blood culture was positive for VREfm on day 213 (Event E, Fig. 1A). Antibiotic therapy was switched to weekly oritavancin infusions, but bacteremia again recurred (Event F, Fig. 1A). A subsequent trial of tigecycline was not tolerated due to GI side effects, and the patient was continued on daptomycin. During this time, all breakthrough BSIs were able to be managed in the outpatient setting in accordance with patient and family preference. However, these BSI recurrences prompted a search for additional phages. On day 303, ΦHi3 (related to a previously characterized siphovirus phage, Φ9183 (5)) was added to the treatment regimen alongside Φ9184 for a total of 2×10^9^ PFU/dose. The patient remained bacteremia-free for 4 months, was able to travel, and was hospitalized only once in the 6 months after starting phage therapy for a non-BSI related reason. IV daptomycin was discontinued after an extended 14-week treatment course. The patient was then trialed on oral suppressive therapy with doxycycline and tedizolid, and the PO and IV phage regimen was eventually decreased to twice daily maintenance therapy. However, on day 395 the patient developed a recurrent *E. faecium* BSI (Event G, Fig. 1A).

Isolates from every BSI event as well as *E. faecium* collected from a stool sample (collected on day 80) and historical epidemiologic surveillance rectal swabs (collected 18 and 6 months prior the first day of observation) underwent WGS on the Illumina platform. A patient-specific, closed reference genome was generated with long-read sequencing and hybrid assembly for the earliest isolate (Methods). Genomic analysis indicated that all BSI isolates were closely related to isolates from the GI tract, in support of our hypothesis that the source of infection was the patient’s GI enterococcal population (Fig S1). To better understand how phage therapy affected the GI enterococcal population, we performed shotgun metagenomics on 100 – 1000 pooled colonies from stool samples collected routinely throughout the observation period that were plated on enterococcal-selective media (Methods). Prior to initiation of phage therapy, the proportion of reads mapping to the *E. faecium* genome and to the VanA operon steadily increased over time and remained dominant despite Φ9184 therapy (Fig. 1B). Treatment with oritavancin, a glycopeptide antibiotic to which the patient was naïve, was associated with a transient decrease in *E. faecium* and VanA operon abundance. However, a subsequent rise in *E. faecium* burden was temporally related to breakthrough BSI event F (Fig. 1B). After a brief period of tigecycline exposure and the addition of ΦHi3, both the *E. faecium* and VanA operon abundance in the GI tract again decreased and remained suppressed for several months. During this time (days 248 – 395) there were no BSI events (Fig. 1A). These data suggest that while the combination of systemic antibiotics and Φ9184 did not durably alter the enterococcal population in the GI tract, administration of tigecycline and/or the addition of ΦHi3 suppressed the *E. faecium* population in the GI tract and prevented BSI recurrence for 147 days.

Despite clinical improvement, the patient had a recurrence of BSI on day 395. Throughout treatment we tested breakthrough isolates to evaluate for emergence of phage resistance, and we monitored serum samples for evidence of phage activity neutralization. All breakthrough BSI isolates remained susceptible to both antibiotics and phages employed (Fig. 2A and Supplemental Table 1). Incubating phages with serum collected before the administration of ΦHi3 resulted in no appreciable decrease in the efficiency of plating (EOP) for either phage. However, serum collected 27 days after starting ΦHi3 (day 330) caused an almost 100-fold reduction in the EOP for both phages. All subsequent serum samples completely neutralized phage activity (Fig. 2B). To further characterize the anti-phage component of the serum neutralization response, we quantified IgG binding to Φ9184 or ΦHi3 lysates using a custom ELISA assay (Methods). Serum collected after neutralization was detected (day 337) showed a significant increase in the half-maximal titers against both Φ9184 and ΦHi3 as compared to serum collected prior to phage therapy (day 181) (Fig. 2C and 2D). These data suggest that administration of the phage cocktail and/or doubling the dose triggered a neutralizing antibody response that inhibited phage activity in the bloodstream and may have contributed to the final breakthrough BSI (event G). As such, after 6 months of treatment, phage therapy was discontinued, and the patient and her family decided to transition to hospice care. The patient continued to experience intermittent *E. faecium* bacteremia until she passed away from pneumonia 7.5 months after discontinuation of phage therapy.

**Figure 2.**
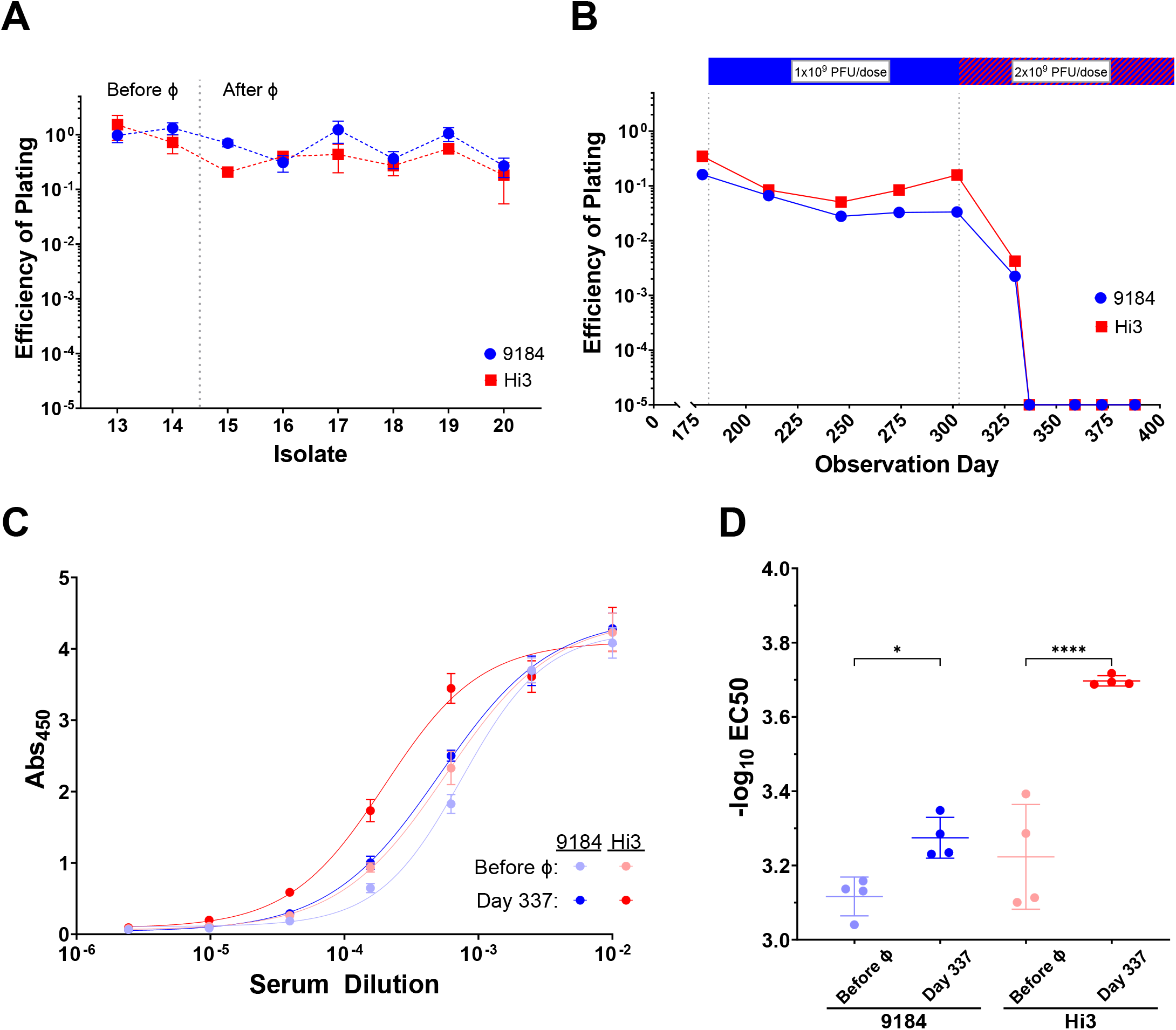
Development of a phage-specific neutralizing immune response. (A) Efficiency of plating (EOP) of Φ9184 and ΦHi3 on clinical *E. faecium* BSI isolates as compared to the phage host strain. Isolates 13 and 14 were the last isolates collected prior to phage initiation (Before Φ), and isolates 15 – 20 represent BSI events occurring after phage therapy was started (After Φ). Error bars show the standard error mean (SEM) between replicates. (B) Average EOP of Φ9184 and ΦHi3 on the host strain after incubation with serum versus incubation with buffer alone, across 3 replicates. (C) ELISA IgG binding curves from host sera against Φ9184 (blue) and ΦHi3 (red) lysates. “Before Φ” indicates the serum sample that was collected prior to initiation of phage therapy (day 181) and “Day 337” indicates serum sample collected on observation day 337, which was 34 days after staring the Φ9184 and ΦHi3 cocktail. Error bars show SEM between replicates. (D) Half-maximal titers (-log_10_ EC50) of each ELISA replicate from the conditions discussed in panel (C) with SEM error bars. * = p < 0.05 and **** = p <0.0001.

*E. faecium* displays resistance to many first-line antibiotics and is regularly associated with persistent and recurrent infections. In this study, systemic antibiotics alone were unable to achieve durable remission of recurrent *E. faecium* BSIs in this patient. A phage cocktail combined with traditional antibiotics did temporarily suppress recurrent BSIs and reduced intestinal *E. faecium* burden, which resulted in fewer hospitalizations and improved quality of life for 6 months. These observations highlight the clinical relevance of prior *in vitro* and pre-clinical studies detailing the synergistic effects of phage cocktails and antibiotic combinations on enterococcal infections (5–7). At this time, there is sparse literature describing clinical experience with *E. faecium-* targeting phages, and few studies have assessed phage-associated changes to gut microbial communities. Additionally, host immune responses are increasingly recognized as a potential limitation of phage therapy (8, 9). To harness the full potential of phages in treating difficult enterococcal infections, future studies should continue to monitor treatment-related microbiome changes and host immune responses to maximize the clinical efficacy of this technology.

## Data Availability

Genome sequencing data for all isolates was submitted to NCBI under BioProject PRJNA901969, with accession numbers listed in Supplemental Table 1. Other data presented are available upon reasonable request to the authors.

## Acknowledgements

We express our deepest thanks to the patient and her family. Without their assistance and perseverance, we would not be able to share our findings with the scientific and medical community. It was the patient’s sincere hope that her experience would inform further studies to help future patients with difficult enterococcal infections. We also thank all members of the Van Tyne lab for helpful input throughout the course of the study, Lee Harrison for providing previously collected bacterial isolates sampled from the patient as part of an ongoing study (R01AI127472), and Marissa Griffith and Vatsala Srinivasa with assistance performing bioinformatic analyses.

Research reported in this publication was supported by the National Institute of Allergy and Infectious Diseases of the National Institutes of Health under Award Number R01AI165519 (to DVT) and R21AI151363 (to RKS). This work was also supported by grants R01AI141479 (BAD), T32AR007534 (MRM), and K23AI154546 (GH) from the NIH. The content is solely the responsibility of the authors and does not represent the official views of the National Institutes of Health. GH is a recipient of research grants from Allovir, Karius, and AstraZeneca. GH also serves on the scientific advisory boards of Karius and AstraZeneca and has received honoraria from MDOutlook. The funders had no role in study design, data collection and analysis, decision to publish, or preparation of the manuscript.

## Supplemental Methods

### Study design

This was a prospective observational case study of a single patient with recurrent *E. faecium* bacteremia that received bacteriophage (phage) therapy. The patient was referred to our laboratory at the request of the attending infectious disease physician and informed patient consent was obtained. FDA emergency investigational new drug approval for administration of phages was obtained (eIND #27183), and the local Institutional Review Board also approved the study (Protocol #EA20120145). The first day of patient observation was in June 2020 and extended through July 2021. Invasive BSI isolates from positive blood cultures were obtained through the UPMC clinical microbiological laboratory and antibiotic minimum inhibitory concentrations (MICs) were measured on the MicroScan WalkAway System (Beckman Coulter) (Supplemental Table 1). Throughout the follow-up period, weekly collection of whole blood and stool samples were planned but were not always available due to outpatient care logistics.

### *E. faecium* isolate whole genome sequencing and assembly

Two VRE isolates from rectal swabs collected from the patient 18- and 6-months prior to the start of observation (isolates labeled REF and PRE, respectively) were available from VRE surveillance efforts that were ongoing during this study. We also collected three clones of *E. faecium* from a stool sample collected from the patient on day 80. Additionally, 20 *E. faecium* isolates collected from BSI events (A – G) were obtained with assistance from the clinical microbiological laboratory.

A total of 25 isolates underwent whole genome sequencing (WGS) on the Illumina platform. Genomic DNA was extracted with a Qiagen DNeasy Blood and Tissue Kit (Qiagen, Hilden, Germany) and Illumina sequencing libraries were made with a Nextera kit (Illumina, San Diego, CA). Libraries were sequenced on a NextSeq 500 using 2×150-bp paired-end reads. The REF (t-18 month) isolate was also sequenced on the Oxford Nanopore Technologies MinION platform, and hybrid assembly was used to generate a patient-specific reference genome with Unicycler(10). Genomes were analyzed using tools available through the Center for Genomic Epidemiology (http://www.genomicepidemiology.org/), and snippy (https://github.com/tseemann/snippy). All isolates were found to belong to multi-locus sequence type (ST) 17 and were separated from the patient-specific reference genome by 18 – 61 single nucleotide polymorphisms (SNPs). A SNP-based, core genome phylogenetic tree was generated with snippy and constructed with RAxML (11) with 100 bootstraps and visualized with iTOL (https://itol.embl.de/login.cgi). The estimated SNP accumulation rate was approximately 18 SNPs/year, which is within previously published ranges for *E. faecium* (12, 13).

### Enterococcal metagenomics

Stool samples were plated onto bile esculin azide agar (BD 212205, Fisher Scientific) and incubated overnight at 37°C. 100 – 1000 colonies were pooled, and genomic DNA was extracted and sequenced on the Illumina platform. Raw reads were assessed for quality (minimum phred quality of 30) and adapters were trimmed with fastp (14). Reads with >90% identity were mapped to the VanA operon with bbmap and the percentage of total reads was calculated (https://sourceforge.net/projects/bbmap/). To determine the species distribution of the pooled enterococcal colonies, a custom database of 1337 enterococcal genomes was constructed from the NCBI GenBank database including all assemblies for Taxon ID 1350 (*Enterococcus)* with Complete Genome assembly level (accessed April 2021). Reads were cleaned as above and all metagenome samples were species classified using Kraken 2 with a confidence score of 0.9 (15). The relative abundance was calculated of the two predominant species, *E. faecalis* and *E. faecium*.

### Phage lysate preparation for treatment

An *E. faecium* isolate from the patient (DVT1320) was used as the host for infection with Φ9184 and ΦHi3. All liquid incubation steps occurred at 37°C and 170 rpm. The host strain was grown overnight to stationary phase in tryptic soy broth (TSB) (Becton Dickinson & Co., BD 211825). The culture was then diluted 1:30 into fresh TSB and CaCl_2_ and MgCl_2_ were each added to a final concentration of 1 mM. Once the culture reached an OD_600_ between 0.15 – 0.25, previously prepared phage lysate was added at multiplicity of infection (MOI) of 0.1. Infected cultures were monitored periodically until the OD_600_ began to decline, signaling phage-mediated bacterial lysis. Cultures were pulled from the incubator and allowed to complete infection overnight at 4°C. The following day, the cultures were cleared with a 15-minute centrifugation at 4000g. The supernatant was filtered through a 22μM polystyrene filter (Corning). The resulting lysate was concentrated roughly 20x using cellulose 30,000 nominal molecular weight limit (NMWL) centrifugal filtration devices (Centricon Plus-70, Merck-Millipore Ltd). Concentrated lysate was then sterilized again by passage through a 22 μM syringe filter (Fisherbrand, cat# 09-720-004) and formulated at the desired titer in 1xPBS with 10mM MgSO_4_. USP71 sterility testing was performed (Accugen Laboratories, Addison, IL) and endotoxin concentration was measured by LAL assay (Thermo Scientific, Waltham, MA).

### Phage spot titer assay

Lytic activity of Φ9184 and ΦHi3 on *E. faecium* clinical isolates was measured using the soft agar overlay method. Briefly, overnight cultures of each isolate were grown at 37°C in tryptic soy broth (TSB) (Becton Dickinson & Co., BD 211825). A 1mL aliquot was pelleted and resuspended in an equal volume of SM+ buffer (50 mM TrisCl pH 7.5, 100 mM NaCl, 8 mM MgSO_4_, 2.5 mM CaCl_2_). Resuspended cultures were diluted 1:50 in 5 mL molten TSB top agar (0.35% Agar, 10 mM MgSO_4_) and layered on top of TSB bottom agar (1.5% Agar, 20 mM MgSO_4_) and allowed to solidify. 10-fold serial dilutions of phage lysates were made in SM+ buffer, and 5 μL of each dilution was spotted onto the top agar. Spots were allowed to dry, and then plates were incubated upright for 24 – 48h at 37°C. The least diluted sample with countable plaques was recorded and titers were calculated as plaque forming units (PFU) per mL of lysate. Efficiency of plating (EOP) was calculated by dividing the phage titer on the clinical isolate by the titer on the host used for phage production.

### Serum neutralization assay

Serum neutralization assays were performed similarly as the spot titer assays above except that 20 μL of each phage lysate was added to 180 μL of patient sera and the mixture was incubated for 24 hours at 37°C. The pre-incubated lysates were then serially 10-fold diluted with SM+ buffer and spotted onto top agar as above. EOP was calculated by dividing the titer of phage incubated with serum by the titer of phage incubated with SM+ buffer alone.

### ELISA

A phage-specific protocol was adapted from Dedrick et al, 2021 and is summarized here (8). 96-well EIA microplates (Corning, CLS3590) were coated with either 100 μL of coating buffer (carbonate-bicarbonate pH 9.6 Sigma-Aldrich, cat# C3041) or 1×10^9^ PFU/mL of phage lysate diluted into coating buffer. Plates were sealed and incubated at 4°C for 20 – 24h. Wells were washed 5 times with 250 μL PBST (PBS – Sigma-Aldrich, P5493; 0.05% Tween-20 – EMD Millipore Corp., 655205), then blocked for 20 – 24h in blocking buffer (PBST + 3% milk – Research Products International, M17200). Patient serum samples were heat inactivated at 56°C for 30 mins, aliquoted and stored at -20°C until use. Starting at an initial dilution of 1:100, Serum samples were serially diluted 1:3 in blocking buffer. Block was removed from wells without washing, and 100 μL of patient serum dilutions were added to each well in technical duplicate. Plates were sealed and incubated at 4°C for 20 – 24h. Wells were then washed 5 times with 250 μL PBST. HRP-conjugated secondary antibody (goat anti-human IgG Fc, Abcam, Cat# ab98624) was diluted 1:10,000 in PBST and 100 μL was added to each well. Plates were sealed and incubated in the dark at room temperature for 1h. Wells were washed 2-3x with 250 μL PBST followed by 2-3x 250 μL PBS. 100 μL of HRP substrate (3,3’,5,5’-tetramethylbenzidine, Sigma, cat# T0440) was added to each well and incubated in the dark at room temperature for 8 mins. The reaction was then stopped by adding 100 μL 2N H_2_SO_4_ and absorbance at 450 nm was measured on a BioTek Synergy H1 plate reader (BioTek, Winooski, VT).

For rows containing wells with absorbance above the limit of detection of the plate reader, the entire row was diluted 1:3 with water in a new plate and absorbance was remeasured. A dilution correction factor was calculated using wells with measurable signal in both undiluted and diluted wells by dividing the signal in the undiluted samples by that of the diluted samples. Diluted well absorbance was then multiplied by the correction factor to obtain the estimated undiluted absorbance for saturated wells. Each serum sample ELISA was performed in duplicate. All replicate values were used to fit the overall curve using a 4-parameter sigmoidal function linear regression as shown in Fig 2C. The same linear regression function was used to fit curves for each individual serum replicate to calculate the individual -logEC50s, which were compared using a one-way ANOVA.

### Statistical Analysis

All statistical analyses and graphing were performed in Graph Pad Prism version 9.4.1. Replicate values were summarized with mean values and standard error means. One-way ANOVA was used to compare between conditions and p-values < 0.05 were considered statistically significant.

### Data Availability

Genome sequencing data for all isolates was submitted to NCBI under BioProject PRJNA901969, with accession numbers listed in Supplemental Table 1.

## Figures

**Supplemental Figure 1.**
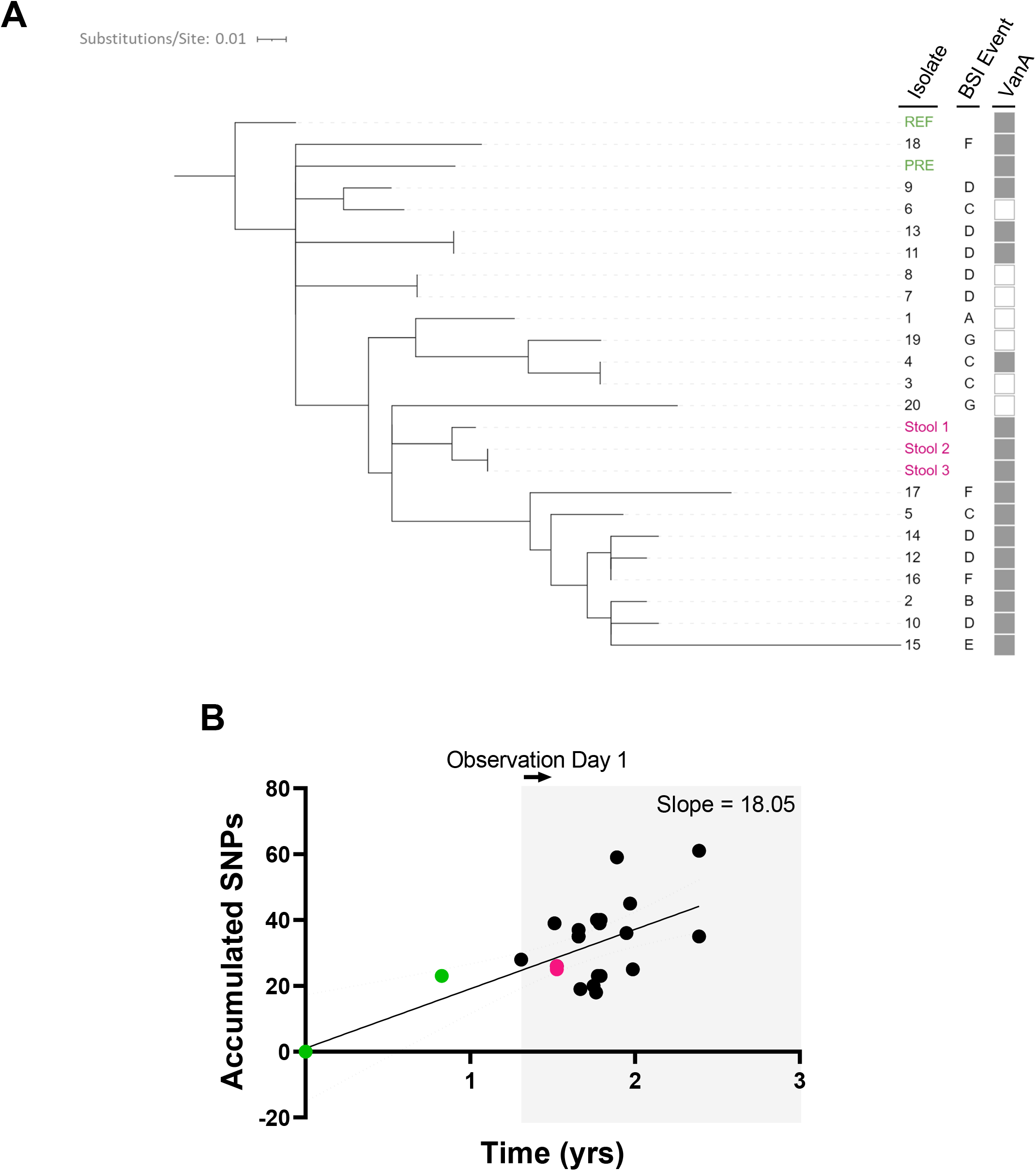
Genomic analysis of recurrent *E. faecium* BSI isolates. (A) Core genome SNP-based maximum likelihood phylogenetic tree built with RAxML with 100 bootstraps. BSI isolate IDs are listed to the right. Pre-observation isolates are in green, invasive BSI isolates are in black and stool sample isolates are in pink. BSI events are listed (A – G, see Fig 1A). Gray color strip indicates presence (shaded) or absence (open) of the VanA operon. (B) Accumulated core genome SNPs over time compared to the reference genome (at origin). Pre-observation, invasive BSI and stool isolates are color-coded as in panel (A). Best fit line estimates the overall SNP accumulation rate of 18 SNPs/year. Dotted curves indicate the 95% confidence bands. The gray background shading indicates the observation period (June 2020 – July 2021).

## References

1. Van Tyne D, Gilmore MS. 2014. Friend turned foe: evolution of enterococcal virulence and antibiotic resistance. Annu Rev Microbiol 68:337–356.

2. Centre for Disease Control. 2019. Antibiotic Resistance Threats in the United States (2019 AR Threats Report). U.S. Department of Health and Human Services, CDC.

3. El Haddad L, Harb CP, Gebara MA, Stibich MA, Chemaly RF. 2019. A Systematic and Critical Review of Bacteriophage Therapy Against Multidrug-resistant ESKAPE Organisms in Humans. Clin Infect Dis 69:167–178.

4. Paul K, Merabishvili M, Hazan R, Christner M, Herden U, Gelman D, Khalifa L, Yerushalmy O, Coppenhagen-Glazer S, Harbauer T, Schulz-Jürgensen S, Rohde H, Fischer L, Aslam S, Rohde C, Nir-Paz R, Pirnay J-P, Singer D, Muntau AC. 2021. Bacteriophage Rescue Therapy of a Vancomycin-Resistant Enterococcus faecium Infection in a One-Year-Old Child following a Third Liver Transplantation. Viruses 13.

5. Canfield GS, Chatterjee A, Espinosa J, Mangalea MR, Sheriff EK, Keidan M, McBride SW, McCollister BD, Hang HC, Duerkop BA. 2021. Lytic bacteriophages facilitate antibiotic sensitization of Enterococcus faecium. Antimicrob Agents Chemother https://doi.org/10.1128/AAC.00143-21.

6. Morrisette T, Lev KL, Kebriaei R, Abdul-Mutakabbir JC, Stamper KC, Morales S, Lehman SM, Canfield GS, Duerkop BA, Arias CA, Rybak MJ. 2020. Bacteriophage-Antibiotic Combinations for Enterococcus faecium with Varying Bacteriophage and Daptomycin Susceptibilities. Antimicrob Agents Chemother 64.

7. Morrisette T, Lev KL, Canfield GS, Duerkop BA, Kebriaei R, Stamper KC, Holger D, Lehman SM, Willcox S, Arias CA, Rybak MJ. 2022. Evaluation of Bacteriophage Cocktails Alone and in Combination with Daptomycin against Daptomycin-Nonsusceptible Enterococcus faecium. Antimicrob Agents Chemother 66:e0162321.

8. Dedrick RM, Freeman KG, Nguyen JA, Bahadirli-Talbott A, Smith BE, Wu AE, Ong AS, Lin CT, Ruppel LC, Parrish NM, Hatfull GF, Cohen KA. 2021. Potent antibody-mediated neutralization limits bacteriophage treatment of a pulmonary Mycobacterium abscessus infection. Nat Med 27:1357–1361.

9. Nick JA, Dedrick RM, Gray AL, Vladar EK, Smith BE, Freeman KG, Malcolm KC, Epperson LE, Hasan NA, Hendrix J, Callahan K, Walton K, Vestal B, Wheeler E, Rysavy NM, Poch K, Caceres S, Lovell VK, Hisert KB, de Moura VC, Chatterjee D, De P, Weakly N, Martiniano SL, Lynch DA, Daley CL, Strong M, Jia F, Hatfull GF, Davidson RM. 2022. Host and pathogen response to bacteriophage engineered against Mycobacterium abscessus lung infection. Cell 185:1860-1874.e12.

10. Wick RR, Judd LM, Gorrie CL, Holt KE. 2017. Unicycler: Resolving bacterial genome assemblies from short and long sequencing reads. PLoS Comput Biol 13:e1005595.

11. Stamatakis A. 2014. RAxML version 8: a tool for phylogenetic analysis and post-analysis of large phylogenies. Bioinformatics 30:1312–1313.

12. Howden BP, Holt KE, Lam MMC, Seemann T, Ballard S, Coombs GW, Tong SYC, Grayson ML, Johnson PDR, Stinear TP. 2013. Genomic insights to control the emergence of vancomycin-resistant enterococci. MBio 4.

13. Duchêne S, Holt KE, Weill F-X, Le Hello S, Hawkey J, Edwards DJ, Fourment M, Holmes EC. 2016. Genome-scale rates of evolutionary change in bacteria. Microb Genom 2:e000094.

14. Chen S, Zhou Y, Chen Y, Gu J. 2018. fastp: an ultra-fast all-in-one FASTQ preprocessor. Bioinformatics 34:i884–i890.

15. Wood DE, Lu J, Langmead B. 2019. Improved metagenomic analysis with Kraken 2. Genome Biol 20:257.

